# Social Determinants of Cost-Related Medication Nonadherence in the All of Us Cohort

**DOI:** 10.1101/2025.05.31.25328711

**Authors:** Tadesse M. Abegaz, Emre Sezgin, Suresh K Bhavnani, Macarius Donneyong

## Abstract

**Background:** This study aimed to examine the social determinants of health (SDH) associated with Cost-related medication nonadherence (CRMNA).

**Methods:** A cross-sectional analysis was conducted using data from the All of Us Research Program to identify SDH features associated with CRMNA. CRMNA include inability to afford prescription medication, skipped doses, reduced dosage, selection of lower-cost alternatives, use of alternative therapies, purchasing medications from another country, and delayed prescription filling. A network analysis using the Fruchterman-Reingold algorithm was performed to visualize correlations among SDH features and CRMNA. SDH variables most strongly correlated with CRMNA were analyzed using binary logistic regression to estimate their association with each CRMNA, adjusting for demographic variables.

**Results:** According to the network analysis, the SDH features most frequently associated with CRMNAs include housing problems, worries about food insecurity, and experiencing poor attention by healthcare professionals. In the adjusted binary logistic regression model, Individuals who reported concerns about food not lasting were more than twice as likely to report overall CRMNA (adjusted odds ratio [AOR] = 2.29; 95% CI: 2.19–2.40; p < 0.05). Other significant SDH predictors of CRMNA included feeling unheard by a doctor (AOR = 1.59; 95% CI: 1.50–1.69; p < 0.05), experiencing housing problems (AOR = 1.40; 95% CI: 1.36–1.46; p < 0.05), and reporting perceived discrimination (AOR = 1.36; 95% CI: 1.32–1.40; p < 0.05).

**Conclusion:** This study highlights the multifaceted impact of SDH on CRMNA, emphasizing the critical roles of food insecurity, housing problem, and negative healthcare experiences in shaping medication adherence behaviors.

## Introduction

Cost-related medication non-adherence (CRMNA) remains a significant public health concern, especially among individuals with chronic illnesses, as it is linked to poorer health outcomes (1). A nationally representative study conducted in 2016 reported that 14.4% of adults aged 65 and older did not adhere to prescribed medications due to cost barriers (2). Similarly, findings from a national panel survey of more than 2,000 participants indicated that approximately one in five Americans experienced CRMNA (3).

Social determinants of health (SDH) refer to the social, economic, and environmental conditions in which people are born, grow, live, work, and age that exert a profound influence on health outcomes across the lifespan (4). Patients experiencing one or more adverse SDH have been shown to be at increased risk for CRMNA. For example, factors such as food insecurity and housing instability have been associated with lower rates of medication adherence (5). A cross-sectional study using data from the *All of Us* (AoU) cohort reported a high prevalence of CRMNA, potentially driven by adverse SDH (6). Another study identified specific SDH, such as unemployment, low income, lower education levels, and residing in states with limited Medicare coverage as independent predictors of CRMNA (7). Similarly, a study examining antihypertensive medication adherence in a Medicaid cohort found that individuals experiencing multiple social adversities demonstrated poorer adherence compared to those facing fewer challenges (8). Specifically, among individuals with low income, the cost of medications has been consistently identified as a significant barrier to adherence (9).

While previous studies have examined the effects of individual SDH on CRMNA, these factors often interact in complex ways and exert varying degrees of influence on adherence behaviors (10). However, the specific SDH features most strongly associated with CRMNA remain insufficiently identified. Moreover, although network analysis methods have been applied to classify patients at risk for adverse outcomes such as depression, emergency department visits, hospital readmission, and COVID-19 infection, their utility in identifying key SDH factors linked to CRMNA has not yet been explored (11, 12) (10). To address this gap, we leveraged the large-scale AoU dataset, which includes over 80 SDH variables from more than 117,000 individuals, and applied the Fruchterman-Reingold network analysis technique. Identifying critical SDH features associated with CRMNA may inform targeted interventions, reduce modifiable risks, and ultimately enhance medication adherence (13).

## Methods

### Study Design

A cross-sectional study was conducted using participants from the AoU cohort who had completed the SDH survey and provided responses to the CRMNA variables in the healthcare utilization survey. As illustrated in Figure 1, the study was conducted in several phases: data extraction, network analysis, regression analysis, and interpretation. During the data extraction phase, participants with complete SDH and CRMNA records were identified. In the second phase, Fruchterman-Reingold network analysis was applied to explore correlations between SDH variables and CRMNA. In the regression phase, binary logistic regression was used to assess the associations between selected SDH features and CRMNA, adjusting for patient demographic characteristics. Finally, during the interpretation phase, the research team met to discuss and contextualize the relationships identified between SDH and CRMNA (Figure 1).

**Figure 1:**
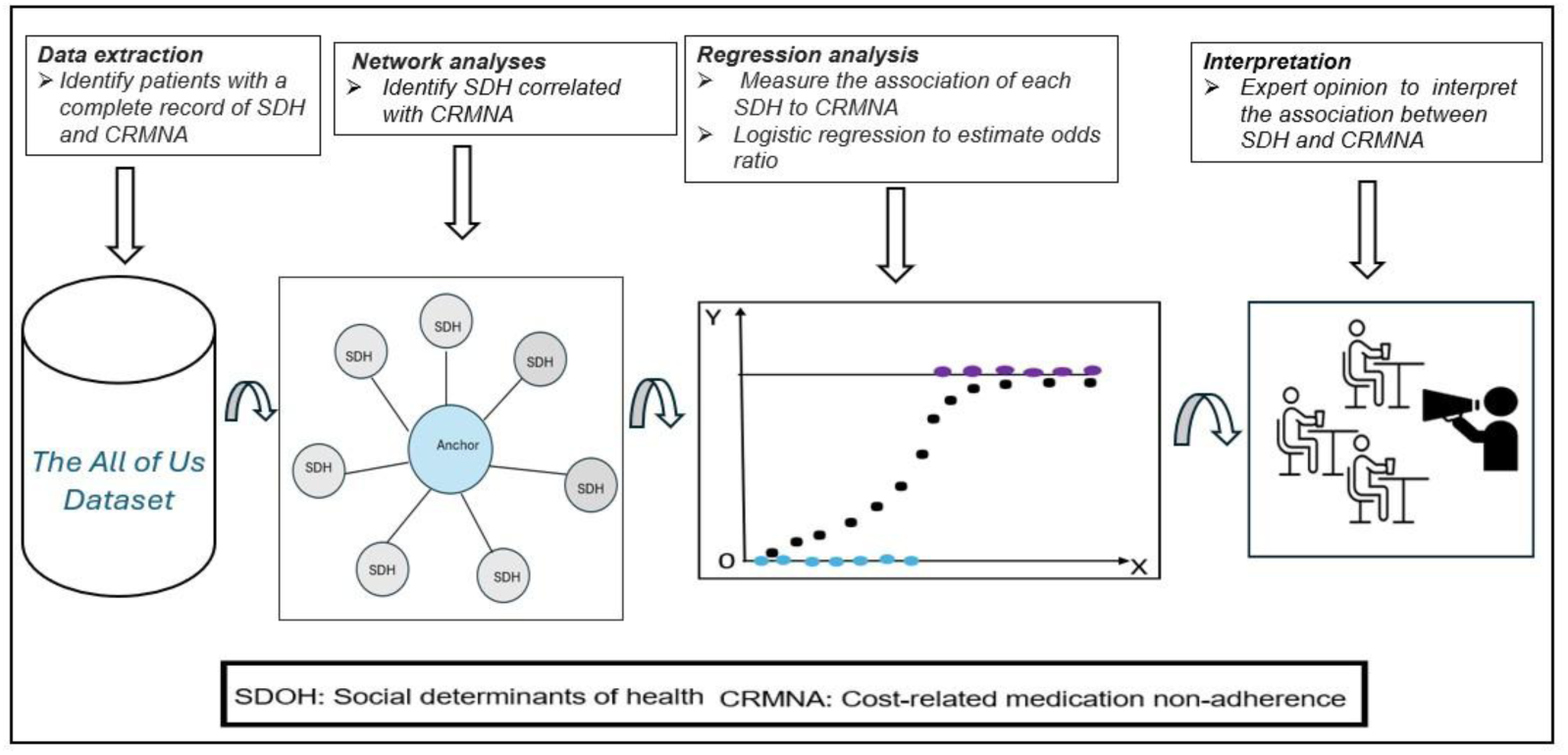
Conceptual framework to examine the association between SDH and CRMNA

### Study Population

All AoU participants aged 18 years and older with complete and valid responses to both the SDH survey and CRMNA items were included in the study.

### Data Sources

Data was sourced from the AoU Research Program. Comprehensive information about the database is available at https://allofus.nih.gov/ (14). In brief, the AoU Research Program, launched in 2018, is a large and diverse cohort comprising participants historically underrepresented in medical research. AoU data includes electronic health records (EHR), survey responses, genomic data, Fitbit data, and physical measurements. The SDH survey component of AoU began on November 1, 2021, with the most recent data collection completed by June 30, 2022. Of all AoU participants, only 117,783 completed the SDH survey, representing a response rate of approximately 30%. CRMNA data, on the other hand, have been collected since the program’s inception as part of the healthcare utilization survey. To ensure alignment with the SDH data collection period (2021–2022), we restricted CRMNA data to the same timeframe.

### Measurements

#### Demographic Characteristics

Sociodemographic characteristics included age, gender, race, ethnicity, income, insurance status, education, and employment. Age was categorized into the following groups: 18–40 years, 40–64 years, 65–74 years, 75–84 years, and 85 years or older (6). Annual household income (in U.S. dollars) was categorized as: ≤$25,000, $25,001–$50,000, $50,001–$100,000, $100,001–$200,000, and >$200,000. Educational attainment was classified into four categories: no high school diploma, high school diploma, some college, and college degree or higher.

#### Social Determinants of Health Features

The AoU dataset organizes SDH features into four major domains: social and community context, economic stability, neighborhood and built environment, and health and healthcare. The social and community context domain includes items related to social cohesion among neighbors (4 items), social support (8 items), loneliness (8 items), perceived discrimination (10 items), perceived stress (10 items), daily spiritual experiences (6 items), religious service attendance (1 item), and English proficiency (1 item). The economic stability domain consists of measures such as food insecurity, a housing instability indicator, and housing quality. The neighborhood and built environment domain comprise neighborhood physical disorder (6 items), neighborhood social disorder (7 items), neighborhood walkability (5 items), neighborhood crime (2 items), and neighborhood residential density (1 item). Lastly, the health and healthcare domain include perceived discrimination in medical care settings (7 items). In this study, over 80 SDH features across these domains were extracted and included in the final analysis. Further details about the SDH variables can be found in the supplementary files (Figure S1, Table S1).

#### Cost-related Medication Nonadherence Variables

The CRMNA was the dependent variable in this study. Participants were asked whether they had engaged in any of the following cost-saving behaviors related to medication adherence in the past year (7 variables): stopped buying medications, skipped doses, reduced dosage, delayed prescriptions, requested cheaper medications, used alternative therapies, or purchased medications from another country. The overall or composite CRMNA score was calculated by summing the responses to these individual questions. CRMNA was considered present if at least one of the above non-adherence behaviors was reported in the past 12 months.

#### Data Extraction Process

The AoU dataset utilizes a cohort builder to facilitate data extraction. The process begins by constructing a cohort of participants based on specified inclusion and exclusion criteria, such as patients with a history of CRMNA and SDH. Three separate datasets were created: the CRMNA dataset, the demographic dataset, and the SDH dataset. The CRMNA dataset included data from the seven CRMNA variables, while the SDH dataset contained data on the SDH variables. The demographic dataset included participant demographic characteristics. These datasets were then merged using the person_id variable as a unique identifier to ensure that all data from the CRMNA, SDH, and demographic datasets were properly linked. Both the SDH and CRMNA variables were recorded as binary variables, where 0 indicated "Yes" (presence of the factor) and 1 indicated "No" (absence of the factor) (Table S2).

### Statistical Analysis

#### Fruchterman-Reingold Network Analysis

The first phase of data analysis involved network analysis to identify SDH features that strongly correlated with the anchor variables, specifically the individual CRMNA variables and the composite CRMNA. In the network, CRMNA variables were treated as "nodes" or "vertices," which were connected by "edges" (ties) to the SDH features. These edges represented the direction and strength of the relationship between the nodes.

To estimate the strength of association between each anchor variable and the SDH features, a correlation matrix was generated using Cramér’s V statistics. Cramér’s V is a measure of association between two categorical variables (15), derived from chi-square statistics. It quantifies the strength of the relationship, with values ranging from 0 (no association) to 1 (perfect association).

The following formula shows how Cramer’s V was calculated from the chi-square statistics.

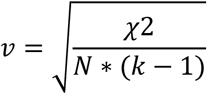

𝑣 stands for Cramer’s V formula/score, where 𝜒^2^ is the chi-square statistic, N is the total sample size of the study population, and k is number of categories/values in the smaller variable in the two-by-two contingency table.

The Cramér’s V values were generated using the VCD package in r, a visualization and modeling package for categorical data (16). The visualization of the network was done with the Fruchterman Reingold layout/algorithm using the igraph package (17). The Fruchterman-Reingold layout is a widely used algorithm for visualizing graphs in network analysis. It arranges nodes and edges in a way that provides minimal overlap between nodes, and clear separation of clusters or groups in the graph. In Fruchterman-Reingold Algorithm clusters of densely connected nodes are positioned close to one another, while sparsely connected nodes are spaced further apart, creating a layout that reflects the graph’s underlying structure.

The CRMNA variables served as anchor variables for the network analysis. To eliminate SDH features with minimal correlation to CRMNA, a unique correlation threshold was established for each anchor variable. For the overall/composite CRMNA (anchor 1), the correlation threshold (v) was set at v = 0.103, which allowed the retrieval of the top 10 SDH features with a strong correlation to the anchor variable. The correlation thresholds for the other CRMNA anchor variables were as follows: 1) skipping medication (anchor 2): v = 0.114, 2) taking alternative therapy (anchor 3): v = 0.09, 3) taking a low dose of the medication (anchor 4): v = 0.11, 4) opting for low-cost medication (anchor 5): v = 0.06, 5) unable to afford prescription (anchor 6): v = 0.12, 6) buying from another country (anchor 7): v = 0.02, and 7) delayed filling (anchor 8): v = 0.27. The top 10 SDH features demonstrating strong correlations with each CRMNA variable were retained for further regression analysis, aimed at exploring their predictive impact on CRMNA, adjusted for patient demographic characteristics.

### Logistic Regression Analysis

The second phase of the analysis involved logistic regression to assess the association between the selected SDH features and the different CRMNA variables. Each CRMNA variable was treated as an outcome variable, with the selected SDH features serving as covariates. Statistical tests were two-sided, and P-values less than 0.05 were considered statistically significant. The analysis was conducted on the NIH All of Us Researcher Workbench using R software version 4.4.0.

## Results

### Sociodemographic Characteristics and Cost-Related Medication Nonadherence

At the time of the study, a total of 413,457 participants were enrolled in the AoU Research Program. Of these, 190,209 completed the CRMNA, and 117,783 responded to the SDH survey. A total of 106,213 participants who provided complete responses to both the CRMNA and SDH surveys were included in the final analysis. Among these participants, 62.58% were female and 77.74% identified as non-Hispanic White. Approximately 34.5% had an education level beyond college, and 26.65% reported an annual income between $50,000 and $100,000.

Overall, 28.6% of participants reported at least one form of CRMNA. Specific CRMNA behaviors included skipping medication (6.9%), using lower-than-prescribed doses (6.8%), opting for alternative therapies (18.1%), being unable to afford prescriptions (9.8%), purchasing medications from another country (3.2%), and delaying prescription filling (9.6%).

When examining CRMNA by gender, females consistently represented the largest proportion across all categories, comprising between 58% and 74% of those reporting CRMNA behaviors. Males accounted for approximately 20% to 38%, with the highest male representation observed in the category of delayed prescription filling (38.5%).

Racial differences were observed. White participants constituted the majority across all CRMNA categories, ranging from approximately 72% to 79.5%. Black participants had lower representation overall but showed relatively higher proportions in the “cannot afford prescription” (13.4%) and “skip medication” (10.5%) categories. With respect to ethnicity, non-Hispanic/Latino individuals accounted for approximately 78% to 88% of CRMNA cases, while Hispanic/Latino participants comprised 11.5% to 21.9%, with their highest representation in the category of purchasing medications from another country (21.9%).

Income level showed a strong inverse relationship with CRMNA. Participants with annual incomes under $25,000 consistently demonstrated the highest prevalence of CRMNA behaviors, ranging from 23% to 40%, with the highest proportions observed in the “cannot afford prescription” (40.2%) and “skip medication” (36.7%) categories. Individuals earning over $200,000 annually accounted for the lowest proportions of CRMNA, ranging from 1.8% to 8.9%. Table 1 below summarizes the sociodemographic characteristics and reported CRMNA behaviors of study participants in the All of Us Research Program (2021–2022).

**Table 1.**
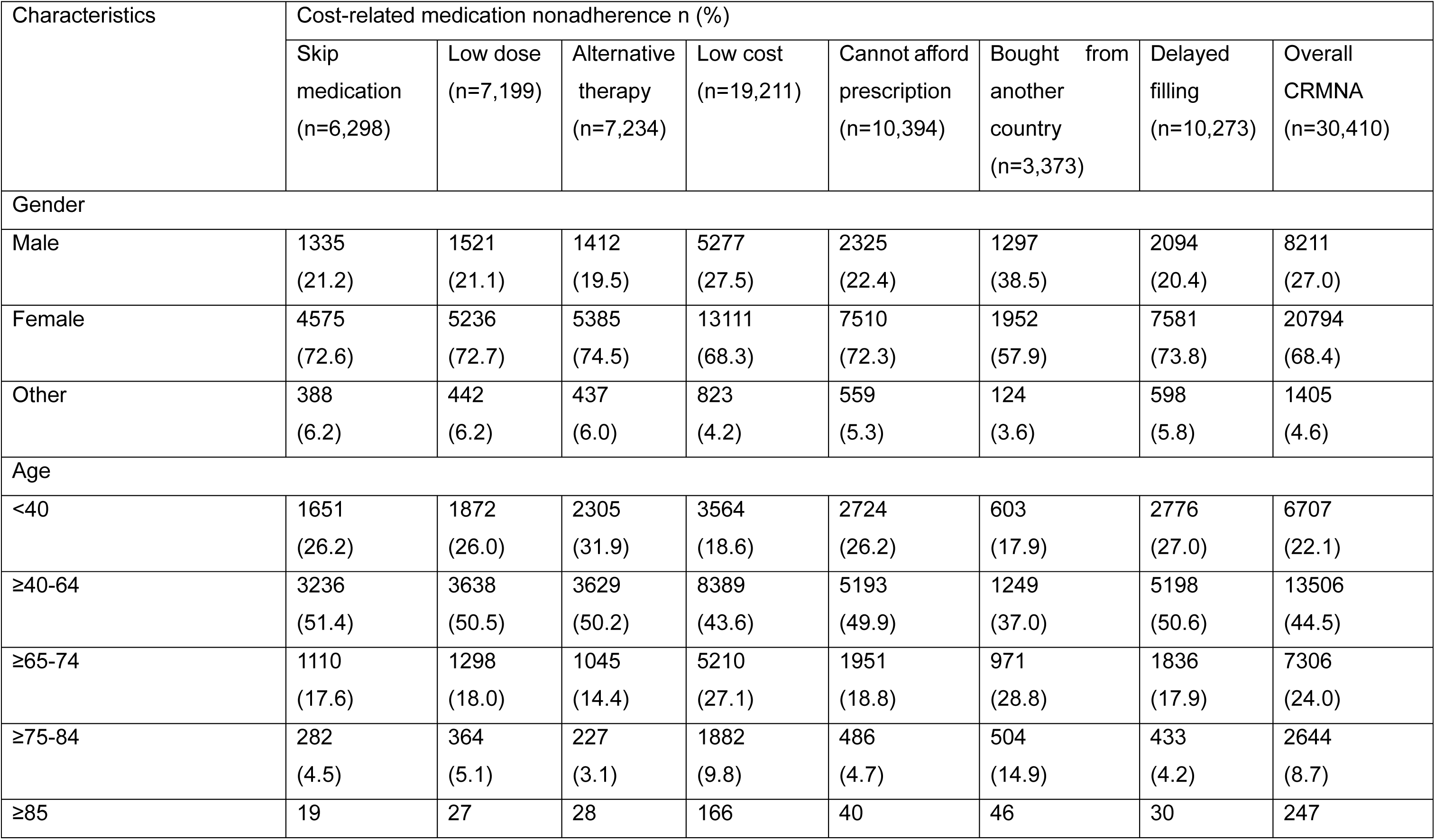
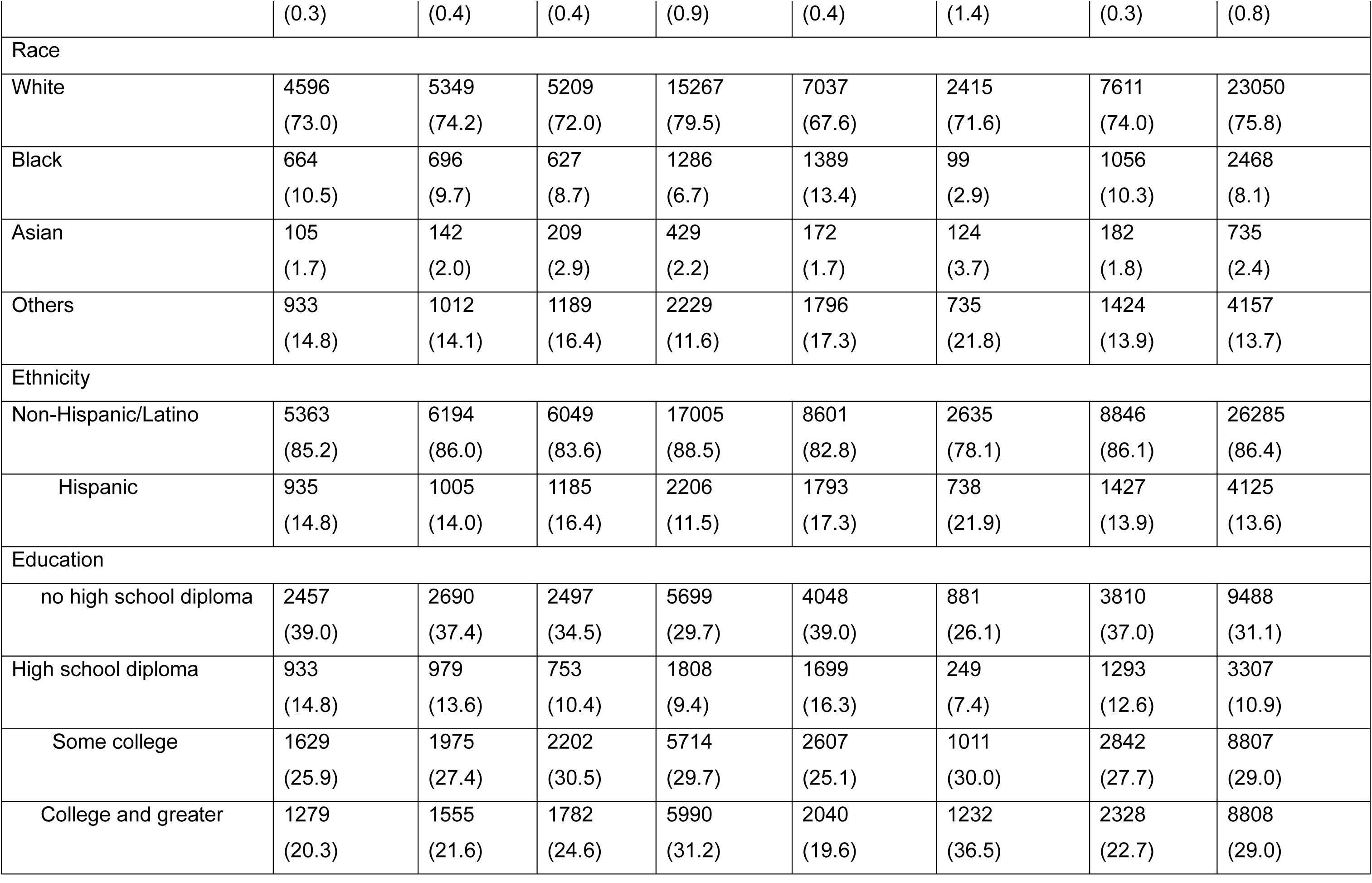
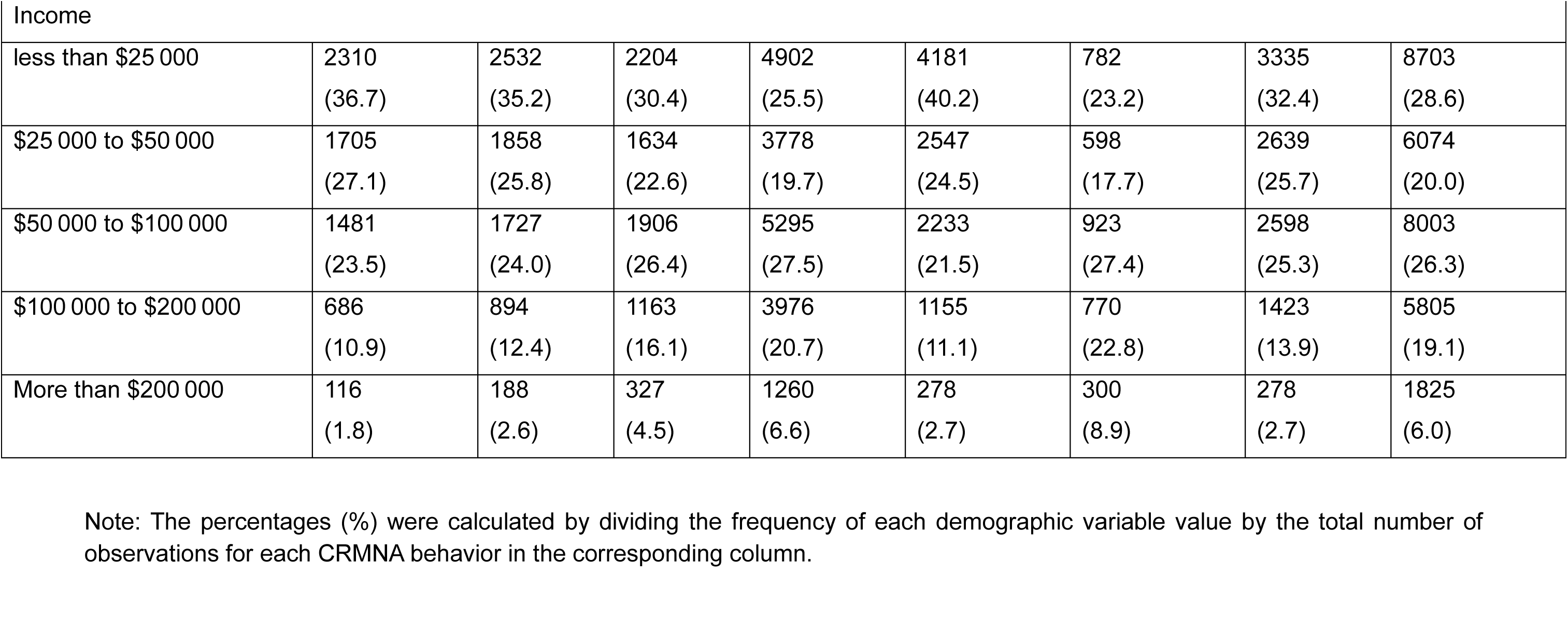
Characteristics of Study Participants and Cost-Related Medication Nonadherence in the AoU Research Program, 2021–2022.

### Fruchterman-Reingold Network and Correlation Coefficients Results

A total of 20 unique SDH features demonstrated strong correlations with all anchor variables, revealing substantial overlap in the SDH features associated with each CRMNA indicator. Among the most frequently linked SDH features were feelings of being treated as inferior, such as experiences where others acted in a superior manner (SDH11) and concerns about life control, including the perception of being unable to control important aspects of one’s life (SDH2).

Food insecurity emerged as a significant theme, particularly concerning food running out before more could be afforded (SDH1, SDH10). Additionally, participants frequently reported negative interactions with healthcare providers, such as being made to feel unintelligent, inferior, or unheard (SDH8, SDH18). Another notable SDH feature included feelings of isolation, such as not having anyone to turn to for suggestions during personal problems (SDH20). A complete list of the 20 SDH features, along with their corresponding domains, is presented in Table 2, while the correlation coefficients for the top ten SDH features are shown in Figure 2.

**Figure 2.**
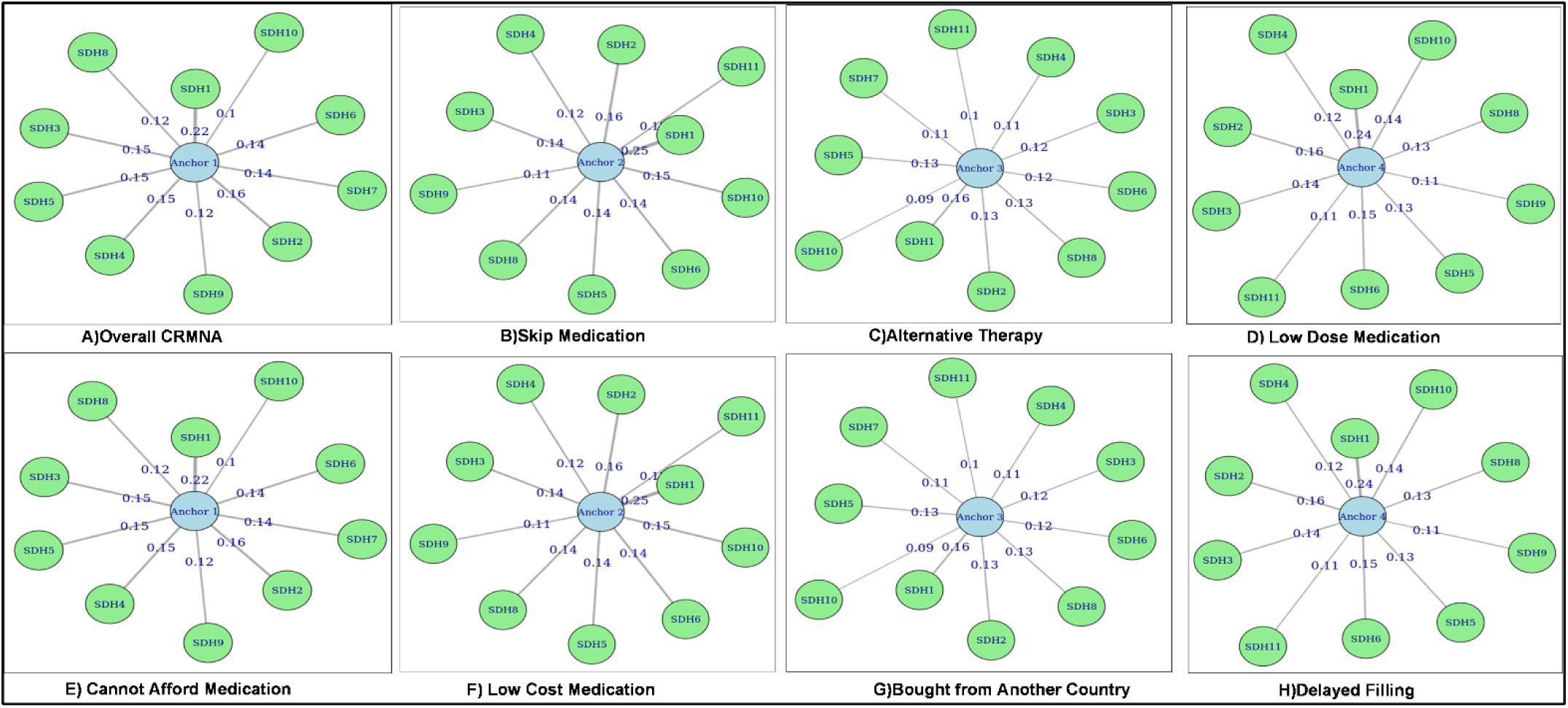
Correlation coefficients between the top ten SDH features and each CRMNA behavior. Each diagram shows an "anchor" node representing a specific CRMNA behavior, with links to the top ten SDH variables. The edge labels indicate the magnitude of the correlation coefficient between the SDH variable and the CRMNA outcome. Thicker or higher-weighted edges represent stronger associations. The SDH features vary slightly across CRMNA behaviors, indicating differential influence of SDH across CRMNA types. Legend: Anchor 1: Overall CRMNA, Anchor 2: Skip medication, Anchor 3: Alternative therapy, Anchor 4: Took less medication/low dose, Anchor 5: Cannot afford prescription, Anchor 6: Low cost, Anchor 7: Buy from another country, Anchor 8: Delayed filling.

**Table 2:** Descriptions of the SDH Features Strongly Correlated with CRMNA Components.

| SDH Code | SDH variable | Domains/measure |
| --- | --- | --- |
| SDH1 | Concern over food running out before you have money to buy more in the past year | Economic stability/food insecurity |
| SDH2 | Feeling unable to control important aspects of your life | Perceived stress |
| SDH3 | Feeling of being on top of things | Perceived Stress |
| SDH4 | Things have felt like they were going your way | Perceived Stress |
| SDH5 | Issues with the place/house where you live | Economic stability/housing stability |
| SDH6 | Being upset because of something that happened unexpectedly | Perceived Stress |
| SDH7 | Discrimination | Perceived discrimination |
| SDH8 | Feeling that healthcare providers are not listening to you | Perceived discrimination |
| SDH9 | Being able to control irritations | Perceived Stress |
| SDH10 | Concern over food running out and lacking money for more in the past year | Economic stability/food insecurity |
| SDH11 | People acting superior to you | Perceived discrimination |
| SDH12 | Language/Speak another language other than English | Health care access |
| SDH13 | Speak English well | Health care access |
| SDH14 | Help with daily chores while sick | Social support |
| SDH15 | Have someone to take you to the doctor if you need it | Social support |
| SDH16 | Have someone to help you if you were confined to bed/ | Social support |
| SDH17 | Have someone who understands your problems | Social support |
| SDH18 | Doctors think they are better than you | Perceived discrimination |
| SDH19 | Have someone to prepare your meals | Social support |
| SDH20 | Have someone for suggestions about how to deal with a personal problem | Social support |

### Association Between SDH Features and Cost-Related Medication Non-Adherence

Binary logistic regression analyses were conducted to evaluate the associations between the top 10 SDH features and both overall and individual CRMNA behaviors, adjusting for demographic characteristics. Individuals who reported concerns about food not lasting (SDH1) were more than twice as likely to report overall CRMNA (adjusted odds ratio [AOR] = 2.29; 95% CI: 2.19–2.40; p < 0.05). Other significant SDH predictors of CRMNA included feeling unheard by a doctor (SDH8) (AOR = 1.59; 95% CI: 1.50–1.69; p < 0.05), experiencing housing problems (SDH5) (AOR = 1.40; 95% CI: 1.36–1.46; p < 0.05), and reporting perceived discrimination (SDH7) (AOR = 1.36; 95% CI: 1.32–1.40; p < 0.05).

For specific CRMNA indicators, similar associations were observed. Participants who reported worrying about food insecurity/SDH1 were significantly more likely to skip medications to save money (AOR = 3.55; 95% CI: 3.33–3.79; p < 0.05), feeling unheard by a doctor/SDH8 (AOR = 1.72; 95% CI: 1.59–1.87; p < 0.05), and report housing problems/SDH5 (AOR = 1.55; 95% CI: 1.46–1.65; p <0.05).

This pattern also extended to the use of alternative therapies, with food insecurity/SDH1 (AOR = 1.82; 95% CI: 1.70–1.94; p < 0.05), feeling unheard/SDH8 (AOR = 1.75; 95% CI: 1.62–1.89; p < 0.05), and housing issues/SDH5 (AOR = 1.59; 95% CI: 1.51–1.68; p < 0.05) all showing significant associations. These three SDH features: food insecurity, communication issues with providers, and housing instability also significantly increased the likelihood of other CRMNA behaviors, including taking a lower dose of medication, opting for low-cost options, delaying prescription fills, or not purchasing medication altogether.

In contrast, different SDH characteristics were associated with purchasing medications outside the U.S. Specifically, perceived provider superiority/SDH18 (AOR = 1.50; 95% CI: 1.30–1.71; *p* < 0.05), limited English proficiency/SDH13 (AOR = 1.24; 95% CI: 1.08–1.43; *p* < 0.05), and lack of support during illness/SDH14 (AOR = 1.19; 95% CI: 1.07–1.33; *p* < 0.05) were all significant predictors of acquiring medications internationally. These findings are illustrated in Figure 3 and Table S3.

**Figure 3:**
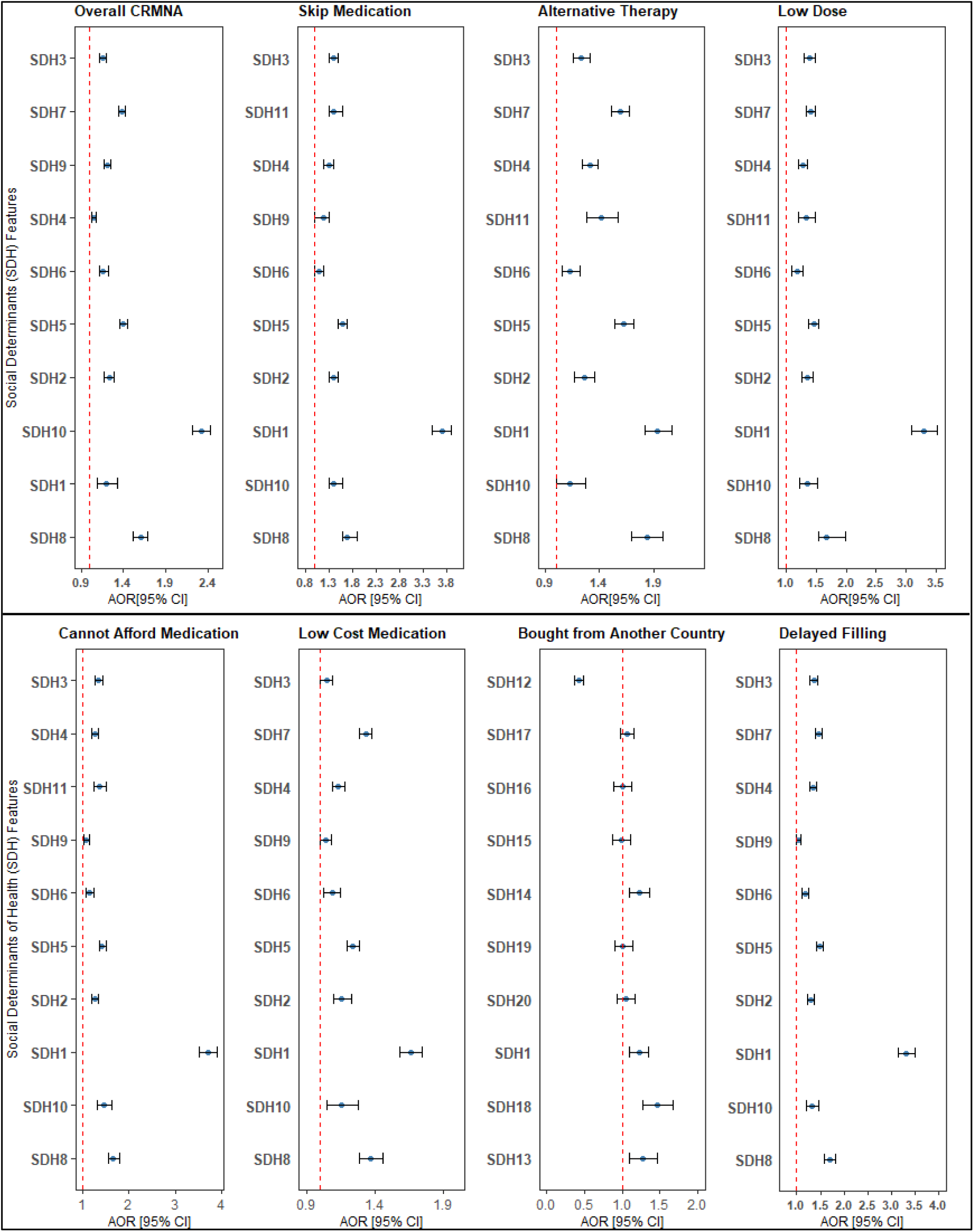
The association between social determinants of health features and each CRMNA

To assess the discriminatory ability of the models, area under the receiver operating characteristic curve (AUC) values were calculated. The AUCs ranged from 0.65 to 0.78 across different CRMNA behaviors, indicating moderate to acceptable discrimination. The model predicting the behavior of purchasing medications from another country yielded an AUC of 0.65, reflecting modest performance. In contrast, the model predicting skipped medication due to cost showed better discrimination, with an AUC of 0.78 (Figure S2).

## Discussion

Social and environmental factors might be related to adverse health outcomes, including medication-nonadherence. This study investigated key social determinants of CRMNA. The primary SDH factors identified as being strongly correlated with CRMNA component include concern about running out of food, housing problems, lack of attention/listening from healthcare providers, discrimination, and inability to control important aspects of life. Among the SDH factors, individuals who reported concerns about food insecurity, such as worrying about running out of food, showed a higher likelihood of medication nonadherence due to cost. Additionally, frequent relocations were associated with a greater probability of nonadherence compared to individuals who remained in stable housing. Furthermore, poor communication and lack of attentiveness from healthcare providers were found to increase the likelihood of nonadherence medication.

The association between medication nonadherence and food insecurity highlights the interconnectedness of basic needs and health behaviors (18). We identified that individuals who experienced food insecurity, which is characterized by concerns about food running out or lasting through the past year, were more likely to report CRMNA. This suggests that financial strain forcing trade-offs between essential needs, such as food and medications, can directly impact adherence behaviors. The relationship between food insecurity and medication non-adherence has been observed in other studies. For instance, a population based cross-sectional study from Canada reported that food-insecure adults have a higher likelihood of cost-related nonadherence to prescription medications than their food-secure counterparts which is consistent with our study findings (19). A meta-analysis study revealed a significant relationship between food insecurity, and housing instability versus medication adherence (5). Food insecurity might force patients to spend their money on food items, get the medication at lower price or delayed filling prescriptions until they have money. In our study, nearly twenty percent of patients reported using low-cost medication and one-tenth reported delaying prescription fills. Therefore, addressing food insecurity as a social determinant of CRMNA may be crucial in mitigating medication nonadherence and improving health outcomes (20).

Furthermore, the present study found that housing instability appeared to be an important social determinant of medication nonadherence. It was associated with an increased risk of nonadherence to medications. Housing instability might be negatively associated with the affordability of care. According to the Downing’s model, housing instability is linked through shifts in healthcare utilization patterns, such as, foregoing medication due to cost which was experienced in 10% of our study population (21). Hernandez et al. indicated that housing insecurity increased the odds of foregoing medication by 64% (22). Therefore, screening of participants for housing instability could reveal potential medication nonadherence issues.

Additionally, the current study found that lack of attentiveness/active listening about patients’ needs by healthcare providers was associated with medication nonadherence. Active listening involves fully focusing on a patient’s concerns and understanding both verbal and nonverbal cues to ensure a comprehensive understanding of their medical needs. This practice is crucial for appropriate care planning, rather than simply issuing prescriptions (23). If communication between the patient and the provider is poor, it is difficult to make shared and informed decision about pharmaceutical care (24). Patients should be involved/engaged in shared decision-making process to improve compliance with recommendations, enhance their understanding of the treatment, and increase knowledge about their medical condition (25). This process will increase the level of agreement or concordance between physician and patient, which has been shown to be associated with improved medication adherence and overall continuity of care (26).

Overall, this study identified the key social determinants of cost-related medication nonadherence using a diverse population who were underserved/underrepresented in medical research. These findings could help inform and extrapolate strategies for addressing nonadherence in these special populations. One of the study’s strengths is the incorporation of a comprehensive list of approximately 80 SDH factors to assess their impact on CRMNA. The study also evaluated all components of CRMNA, such as low-dose prescriptions, low-cost alternatives, skipping doses, delayed prescription filling, enabling an examination of how SDH factors interact with various CRMNAs. The application of a novel approach, the Fruchterman-Reingold network analysis, to reduce data dimensionality while retaining important features strongly correlated with CRMNA highlights methodological strength of the study (27). Although the correlation coefficients generated through the network analysis were relatively lower, the association between SDH features and CRMNA was further corroborated by the logistic regression analysis. Additionally, the transformation of the SDH features that were rated in Likert-scale led to the generation of a more interpretable/meaningful binary variable which could enhance the utility of these features in predicting the risk of CRMNA. Alternatively, newly released scoring methods for evaluating participant responses on SDH features can be considered/implemented in the future for reproducibility of the findings (28).

While this study highlights important social determinants of medication nonadherence, certain limitations should be considered when interpreting the findings. The cross-sectional design precludes causal inferences (29). Additionally, the binary transformation of Likert-scale variables, while enhancing interpretability, may oversimplify complex relationships, and relatively low correlation coefficients from the Fruchterman-Reingold network analysis warrant cautious interpretation of associations. Unmeasured confounders, such as income and employment status, may also play a role in CRMNA, highlighting the need for future research to explore these factors. Despite these limitations, the study’s inclusion of a diverse population, comprehensive assessment of SDH factors, and novel methodological approaches provide valuable insights into addressing CRMNA, particularly in underserved populations.

## Conclusion

This cross-sectional analysis highlights the substantial impact of SDH on cost-related medication non-adherence. Key SDH factors including food insecurity, feeling unheard by healthcare providers, housing instability, and experiences of discrimination—were significantly associated with both overall and specific types of CRMNA. Healthcare providers are encouraged to screen these SDH indicators, as they may identify patients at elevated risk of financially driven medication non-adherence. Early identification of individuals facing adverse SDH can support timely, targeted interventions to prevent non-adherence as a coping strategy for unaffordable medications.

Building strong patient-provider relationships, grounded in empathy and trust, is vital to addressing CRMNA and improving medication adherence.

## Data Availability

All data produced in the present study are available upon reasonable request to the authors

## Acknowledgements

The authors would like to thank The Ohio State University for its overall support. We also extend our gratitude to the *All of Us* Research Program for providing access to the data used in this study. “The All of Us Research Program is supported by the National Institutes of Health, Office of the Director: Regional Medical Centers: 1 OT2 OD026549; 1 OT2 OD026554; 1 OT2 OD026557; 1 OT2 OD026556; 1 OT2 OD026550; 1 OT2 OD 026552; 1 OT2 OD026553; 1 OT2 OD026548; 1 OT2 OD026551; 1 OT2 OD026555; IAA #: AOD 16037; Federally Qualified Health Centers: HHSN 263201600085U; Data and Research Center: 5 U2C OD023196; Biobank: 1 U24 OD023121; The Participant Center: U24 OD023176; Participant Technology Systems Center: 1 U24 OD023163; Communications and Engagement: 3 OT2 OD023205; 3 OT2 OD023206; and Community Partners: 1 OT2 OD025277; 3 OT2 OD025315; 1 OT2 OD025337; 1 OT2 OD025276. In addition, the All of Us Research Program would not be possible without the partnership of its participants.”

## Data availability

The authors confirm that the data supporting the findings of this study are available within the article and its supplementary materials.

## Ethics statement

The *All of Us* Research Program is approved by the *All of Us* Institutional Review Board (IRB). The current study used only de-identified data accessed through the *All of Us* Researcher Workbench and was therefore deemed exempt from additional IRB review.

## Informed consent

Informed consent was not required for this study, as it utilized de-identified secondary data from the *All of Us* Research Program.

## Author contributions

Conceptualization, T.M.A., A.B., and M.D.; Data curation, T.M.A.; Formal analysis, T.M.A.; Investigation, T.M.A.; Methodology, T.M.A., Software, T.M.A; Supervision, M.D.; Validation, T.M., and M.D; Visualization, T.M.A.; Writing—original draft, T.M.A; Writing—review and editing, T.M.A, and M.D. Both authors have read and agreed to the published version of the manuscript.

**Appendix A. Supplementary data**

## References

1. Van Alsten SC. Cost-related nonadherence and mortality in patients with chronic disease: a multiyear investigation, National Health Interview Survey, 2000–2014. Preventing Chronic Disease. 2020;17.

2. Nekui F, Galbraith AA, Briesacher BA, Zhang F, Soumerai SB, Ross-Degnan D, et al. Cost-related medication nonadherence and its risk factors among Medicare beneficiaries. Medical care. 2021;59(1):13–21.

3. Dusetzina SB, Besaw RJ, Whitmore CC, Mattingly TJ, Sinaiko AD, Keating NL, et al. Cost-related medication nonadherence and desire for medication cost information among adults aged 65 years and older in the US in 2022. JAMA Network Open. 2023;6(5):e2314211-e.

4. 4. National Cancer Institute. Social Determinants of Health. https://www.cancer.gov/publications/dictionaries/cancer-terms/def/social-determinants-of-health. Accessed on 19 August 20024.

5. Wilder ME, Kulie P, Jensen C, Levett P, Blanchard J, Dominguez LW, et al. The impact of social determinants of health on medication adherence: a systematic review and meta-analysis. Journal of general internal medicine. 2021;36:1359–70.

6. Delavar A, Saseendrakumar BR, Weinreb RN, Baxter SL. Racial and ethnic disparities in cost-related barriers to medication adherence among patients with glaucoma enrolled in the National Institutes of Health All of Us Research Program. JAMA ophthalmology. 2022;140(4):354–61.

7. Kherallah R, Al Rifai M, Kamat I, Krittanawong C, Mahtta D, Lee MT, et al. Prevalence and predictors of cost-related medication nonadherence in individuals with cardiovascular disease: Results from the Behavioral Risk Factor Surveillance System (BRFSS) survey. Preventive medicine. 2021;153:106715.

8. Wilder ME, Zheng Z, Zeger SL, Elmi A, Katz RJ, Li Y, et al. Relationship between social determinants of health and antihypertensive medication adherence in a Medicaid cohort. Circulation: Cardiovascular Quality and Outcomes. 2022;15(2):e008150.

9. Rohatgi KW, Humble S, McQueen A, Hunleth JM, Chang S-H, Herrick CJ, et al. Medication adherence and characteristics of patients who spend less on basic needs to afford medications. The Journal of the American Board of Family Medicine. 2021;34(3):561–70.

10. Bhavnani SK, Zhang W, Bao D, Raji M, Ajewole V, Hunter R, et al. Subtyping Social Determinants of Health in All of Us: Network Analysis and Visualization Approach. Medrxiv. 2023.

11. Bhavnani SK, Kummerfeld E, Zhang W, Kuo Y-F, Garg N, Visweswaran S, et al. Heterogeneity in COVID-19 patients at multiple levels of granularity: from biclusters to clinical interventions. AMIA Summits on Translational Science Proceedings. 2021;2021:112.

12. Bhavnani SK, Zhang W, Visweswaran S, Raji M, Kuo Y-F. A Framework for Modeling and Interpreting Patient Subgroups Applied to Hospital Readmission: Visual Analytical Approach. JMIR Medical Informatics. 2022;10(12):e37239.

13. 13. The office of disease prevention and health promotion. Social Determinants of Health. https://health.gov/healthypeople/priority-areas/social-determinants-health. Accessed on 19 August 2024.

14. 14. National Institutes of Helath. The All of Us Research Program. https://allofus.nih.gov/. Accesssed 20 August 2024

15. Cramér H. Mathematical Methods of Statistics. Princeton (NJ): Princeton University Press; 1946.

16. Friendly M, Meyer D. Discrete data analysis with R: visualization and modeling techniques for categorical and count data: CRC Press; 2015.

17. Fruchterman TM, Reingold EM. Graph drawing by force-directed placement. Software: Practice and experience. 1991;21(11):1129–64.

18. Ogungbe O, Ellis A, Ojinnaka CO. Food Security to Medication Adherence—Connecting Needs. JAMA network open. 2024;7(2):e2356570-e.

19. Men F, Gundersen C, Urquia ML, Tarasuk V. Prescription medication nonadherence associated with food insecurity: a population-based cross-sectional study. Canadian Medical Association Open Access Journal. 2019;7(3):E590–E7.

20. O’Connor EA, Webber EM, Martin AM, Henninger ML, Eder ML, Lin JS. Preventive services for food insecurity: evidence report and systematic review for the US Preventive Services Task Force. JAMA. 2025.

21. Downing J. The health effects of the foreclosure crisis and unaffordable housing: a systematic review and explanation of evidence. Social Science & Medicine. 2016;162:88–96.

22. Hernandez M, Wong R, Yu X, Mehta N. In the wake of a crisis: Caught between housing and healthcare. SSM-Population Health. 2023;23:101453.

23. Epstein RM, Beach MC. “I don’t need your pills, I need your attention:” Steps toward deep listening in medical encounters. Current Opinion in Psychology. 2023:101685.

24. Bezreh T, Laws MB, Taubin T, Rifkin DE, Wilson IB. Challenges to physician–patient communication about medication use: a window into the skeptical patient’s world. Patient preference and adherence. 2011:11–8.

25. Krist AH, Tong ST, Aycock RA, Longo DR. Engaging patients in decision-making and behavior change to promote prevention. Information Services & Use. 2017;37(2):105–22.

26. Kerse N, Buetow S, Mainous AG, Young G, Coster G, Arroll B. Physician-patient relationship and medication compliance: a primary care investigation. The Annals of Family Medicine. 2004;2(5):455–61.

27. Niu F, Zhao X, Guo J, Shi M, Liu X, Liu B. Fast and robust unsupervised dimensionality reduction with adaptive bipartite graphs. Knowledge-Based Systems. 2023;276:110680.

28. Koleck TA, Dreisbach C, Zhang C, Grayson S, Lor M, Deng Z, et al. User guide for social determinants of health survey data in the All of Us Research Program. Journal of the American Medical Informatics Association. 2024;31(12):3032–41.

29. Larson RB. Controlling social desirability bias. International Journal of Market Research. 2019;61(5):534–47.

