## Supplemental Data 1 for "Social Determinants of Cost-Related Medication Nonadherence in the All of Us Cohort"

*Macarius Donneyong

(614) 292-0075

Table S1: Description of SDH constructs reported in the All of Us research program

Table S2: Relabeling of the SDH and CRMNA Variables

Table S3: Association between SDH features and CRMNA

Figure S1: Illustration of the domains of SDH in the All of Us Research Program

Figure S2: ROC curves for predicting cost-related medication non-adherence

Table S1: Description of SDH constructs reported in the All of Us research program

| SDH Construct | Definitions |
|---|---|
| Social cohesion among neighbors (4-items) | A measure of connection and trust among neighbors, and people's willingness to intervene for the common good |
| Social support (8-items) | A measure of functional, and emotional /informational support in community |
| Loneliness (8-items) | Indicates the severity of feelings of being isolated |
| Perceived discrimination (10-items) | A measure of how often individual are treated with courtesy |
| Perceived stress (10-items) | An instrument which measures how stressful are situations for individuals |
| Daily spiritual experiences (6-items) | Self-report measure of religiousness and spirituality |
| Religious service attendance (1-item) | Assesses how often do people go to religious meetings or services |
| English proficiency (1-item) | It indicates how well individuals speak English |
| Food insecurity (2-item) | It denotes worry whether food would run out |
| Housing instability (1-item) | It shows how often people moved within 1 year |
| Housing quality problem (1-item) | It indicates poor house standards such as infestation, and leaking |
| Neighborhood physical disorder (6-item) | Features of neighborhoods that may signal the breakdown of order |
| Neighborhood social disorder (7-item) | Social features of neighborhoods indicate lack of social control |
| Neighborhood walkability (5-item) | Indicates a walkable neighborhood with high population density, and multimodal transportation |
| Neighborhood crime (2-item) | It refers burglary, vehicle-related crime, and theft |
| Neighborhood residential density (1-item) | The number of people within a geographic area that will support transit |
| Perceived discrimination in medical care settings (7-item) | Perception negative treatment because of one's membership in a particular demographic group (7) |

| | |
|---|---|
| Education (1-item) | The highest grade or year of school you completed |
| Income (1-item) | Annual household income from all sources |
| Employment (1-item) | Indicates current employment status |
| Home ownership/ homelessness (1-item) | Inquiry if individuals own or rent the place where they live |

Table S2: Relabeling of the SDH and CRMNA Variables

| Variable code | Question | Original response | Reclassification/new response | Numerical code |
|---|---|---|---|---|
| discrimination | What do you think is the main reason for these experiences | Reasons include age, gender, race--- | Yes, if mentioned any reason, otherwise no | 0 if yes, 1 otherwise |
| language | Do you speak a language other than English at home | Yes or no | Yes, if one can speak English, otherwise no | 0 if yes, 1 otherwise |
| watchout | How much you agree or disagree that in your neighborhood people watch out for each other | ● Strongly disagree<br>● Disagree<br>● Agree<br>● Strongly agree | Yes, if agree or strongly agree, no, otherwise | 1 if yes, 0 otherwise |
| neighber_help | How much you agree or disagree that people around here are willing to help their neighbor? | ● Strongly disagree<br>● Disagree<br>● Agree<br>● Strongly agree | Yes, if agree or strongly agree, no, otherwise | 1 if yes,0 otherwise |
| trusted_neighber | How much you agree or disagree that people in your neighborhood can be trusted? | ● Strongly disagree<br>● Disagree<br>● Agree<br>● Strongly agree | Yes, if agree or strongly agree, no, otherwise | 1 if yes,0 otherwise |
| get_along | 'How much you agree or disagree that people in your neighborhood generally get along with each other? | ● Strongly disagree<br>● Disagree<br>● Agree<br>● Strongly agree | Yes, if agree or strongly agree, no, otherwise | 1 if yes,0 otherwise |
| share_value | How much you agree or disagree that people in your neighborhood share the same values? | ● Strongly disagree<br>● Disagree<br>● Agree<br>● Strongly agree | Yes, if agree or strongly agree, no, otherwise | 1 if yes,0 otherwise |
| house_care | How much you agree or disagree that people in your neighborhood take good care of their houses and apartments? | ● Strongly disagree<br>● Disagree<br>● Agree<br>● Strongly agree | Yes, if agree or strongly agree, no, otherwise | 1 if yes,0 otherwise |

| | | | | |
|---|---|---|---|---|
| abandoned_buildings | How much you agree or disagree that there are lot of abandoned buildings in your neighborhood? | ● Strongly disagree<br>● Disagree<br>● Agree<br>● Strongly agree | Yes, if agree or strongly agree, no, otherwise | 0 if yes,1 otherwise |
| hang_around | How much you agree or disagree that there are too many people hanging around on the streets near your home? | ● Strongly disagree<br>● Disagree<br>● Agree<br>● Strongly agree | Yes, if agree or strongly agree, no, otherwise | 0 if yes,1 otherwise |
| crime | How much you agree or disagree that there is a lot of crime in your neighborhood? | ● Strongly disagree<br>● Disagree<br>● Agree<br>● Strongly agree | Yes, if agree or strongly agree, no, otherwise | 0 if yes,1 otherwise |
| graffiti | How much you agree or disagree that there is a lot of graffiti in your neighborhood | ● Strongly disagree<br>● Disagree<br>● Agree<br>● Strongly agree | Yes, if agree or strongly agree, no, otherwise | 0 if yes,1 otherwise |
| alcohol | How much you agree or disagree that there is too much alcohol use in your neighborhood? | ● Strongly disagree<br>● Disagree<br>● Agree<br>● Strongly agree | Yes, if agree or strongly agree, no, otherwise | 0 if yes,1 otherwise |
| drug_use | How much you agree or disagree that there is too much drug use in your neighborhood? | ● Strongly disagree<br>● Disagree<br>● Agree<br>● Strongly agree | Yes, if agree or strongly agree, no, otherwise | 0 if yes,1 otherwise |
| vandalism | How much you agree or disagree that vandalism is common in your neighborhood? | ● Strongly disagree<br>● Disagree<br>● Agree<br>● Strongly agree | Yes, if agree or strongly agree, no, otherwise | 0 if yes,1 otherwise |
| trouble_with_neighbors | How much you agree or disagree that you are always having trouble with your neighbors? | ● Strongly disagree<br>● Disagree<br>● Agree<br>● Strongly agree | Yes, if agree or strongly agree, no, otherwise | 0 if yes,1 otherwise |

| | | | | |
|---|---|---|---|---|
| clean_neighbohood | How much you agree or disagree that your neighborhood is clean? | ● Strongly disagree<br>● Disagree<br>● Agree<br>● Strongly agree | Yes, if agree or strongly agree, no, otherwise | 0 if yes,1 otherwise |
| noisy | How much you agree or disagree that your neighborhood is noisy? | ● Strongly disagree<br>● Disagree<br>● Agree<br>● Strongly agree | Yes, if agree or strongly agree, no, otherwise | 0 if yes,1 otherwise |
| safe_neighborhood | How much you agree or disagree that your neighborhood is safe? | ● Strongly disagree<br>● Disagree<br>● Agree<br>● Strongly agree | Yes, if agree or strongly agree, no, otherwise | 0 if yes,1 otherwise |
| less_courtesy_doctor | How often are you treated with less courtesy than other people when you go to a doctor's office or other health care provider? | ● Never<br>● Rarely<br>● Sometimes<br>● Most of the time<br>● Always | yes, if most of the time or always, no, otherwise | 0 if yes, 1, otherwise |
| ess_respect_doctor | How often are you treated with less respect than other people when you go to a doctor's office or other health care provider? | ● Never<br>● Rarely<br>● Sometimes<br>● Most of the time<br>● Always | yes, if most of the time or always, no, otherwise | 0 if yes, 1, otherwise |
| close_toGod | How often do you desire to be closer to or in union with God (or a higher power)? | ● Many times a day<br>● Every day<br>● Most days<br>● Some days<br>● Once in a while<br>● Never or almost never<br>● I do not believe in God (or a higher power) | Yes, if many times a day, every day or most days, no, otherwise | 1 if yes, otherwise 0 |
| gods_love | How often do you feel God's (or a higher power's) love for you, directly or through others? | ● Many times a day<br>● Every day<br>● Most days | Yes, if many times a day, every day or | 1 if yes, otherwise 0 |

| | | | | |
|---|---|---|---|---|
| | | ● Some days<br>● Once in a while<br>● Never or almost never<br>● I do not believe in God (or a higher power) | most days, no, otherwise | |
| gods_presence | How often do you feel God's (or a higher power's) presence? | ● Many times a day<br>● Every day<br>● Most days<br>● Some days<br>● Once in a while<br>● Never or almost never<br>● I do not believe in God (or a higher power) | Yes, if many times a day, every day, or most days, no, otherwise | 1 if yes, otherwise 0 |
| deep_peace | How often do you feel deep inner peace or harmony? | ● Many times a day<br>● Every day<br>● Most days<br>● Some days<br>● Once in a while<br>● Never or almost never<br>● I do not believe in God (or a higher power) | Yes, if many times a day, every day and most days, no, otherwise | 1 if yes, otherwise 0 |
| comfort_religion | How often do you find strength and comfort in your religion? | ● Many times a day<br>● Every day<br>● Most days<br>● Some days<br>● Once in a while<br>● Never or almost never<br>● I do not believe in God (or a higher power) | Yes, if many times a day, every day and most days, no, otherwise | 1 if yes, otherwise 0 |
| spiritually_touched | How often do you feel that you are spiritually touched by the beauty of creation? | ● Many times, a day<br>● Every day<br>● Most days<br>● Some days | Yes, if many times a day, every day and most days, no, otherwise | 1 if yes, otherwise 0 |

| | | | | |
|---|---|---|---|---|
| | | ● Once in a while<br>● Never or almost never<br>● I do not believe in God (or a higher power) | | |
| religious_service | How often do you go to religious meetings or services? | ● More than once a week<br>● Once a week<br>● 1 to 3 times per month<br>● Less than once per month<br>● Never (or almost never)<br>● I am not religious | No, if never (or almost never) or I am not religious, otherwise, yes | 0 if no, otherwise 1 |
| Isolated | How often do you feel isolated from others? | ● Never<br>● Rarely<br>● Sometimes<br>● Often | Yes, if often, no, otherwise | 0 if yes, otherwise,1 |
| lack_campanionship | How often do you feel lack companionship? | ● Never<br>● Rarely<br>● Sometimes<br>● Often | Yes, if often, no, otherwise | 0 if yes, otherwise,1 |
| left_out | How often do you feel left out? | ● Never<br>● Rarely<br>● Sometimes<br>● Often | Yes, if often, no, otherwise | 0 if yes, otherwise,1 |
| doctor_nolistening | How often do you feel like a doctor or nurse is not listening to what you were saying. when you go to a doctor's office or other health care provider? | ● Never<br>● Rarely<br>● Sometimes<br>● Often | Yes, if often, no, otherwise | 0 if yes, otherwise,1 |
| people_notwithyou | How often do you feel that people are around you but not with you? | ● Never<br>● Rarely<br>● Sometimes<br>● Often | Yes, if often, no, otherwise | 0 if yes, otherwise,1 |

| | | | | |
|---|---|---|---|---|
| turn_to | How often do you feel that there is no one you can turn to? | ● None of the time<br>● A little of the time<br>● Some of the time<br>● Most of the time<br>● All of the time | Yes, if Most of the time or All of the time | 0 if yes, otherwise,1 |
| outgoing_person | How often do you feel that you are an outgoing person? | ● Never<br>● Rarely<br>● Sometimes<br>● Often | Yes, if often, otherwise no | 1 if yes, otherwise,0 |
| withdrawn | How often do you feel that you are unhappy being so withdrawn? | ● Never<br>● Rarely<br>● Sometimes<br>● Often | Yes, if often, otherwise no | 0 if yes, otherwise,1 |
| good_time | How often do you have someone to have a good time with? | ● None of the time<br>● A little of the time<br>● Some of the time<br>● Most of the time<br>● All of the time | Yes, if most of the time or all of the time, no, otherwise | 1 if yes, otherwise,0 |
| confined_bed | How often do you have someone to help you if you were confined to bed? | ● None of the time<br>● A little of the time<br>● Some of the time<br>● Most of the time<br>● All of the time | Yes, if most of the time or all of the time, no, otherwise | 1 if yes, otherwise,0 |
| daily_chores | How often do you have someone to help you with daily chores if you were sick? | ● None of the time<br>● A little of the time<br>● Some of the time<br>● Most of the time<br>● All of the time | Yes, if most of the time or All of the time, no, otherwise | 1 if yes, otherwise,0 |
| feel_wanted | How often do you have someone to love and make you feel wanted? | ● None of the time<br>● A little of the time<br>● Some of the time<br>● Most of the time<br>● All of the time | Yes, if most of the time or all of the time, no, otherwise | 1 if yes, otherwise,0 |

| | | | | |
|---|---|---|---|---|
| prepare_meal | How often do you have someone to prepare your meals if you were unable to do it yourself? | ● None of the time<br>● A little of the time<br>● Some of the time<br>● Most of the time<br>● All of the time | Yes, if most of the time or all of the time, no, otherwise | 1 if yes, otherwise,0 |
| take_doctor | How often do you have someone to take you to the doctor if you need it?' | ● None of the time<br>● A little of the time<br>● Some of the time<br>● Most of the time<br>● All of the time | Yes, if most of the time or All of the time, no, otherwise | 1 if yes, otherwise,0 |
| personal_problem_sugestion | How often do you have someone to turn to for suggestions about how to deal with a personal problem? | ● None of the time<br>● A little of the time<br>● Some of the time<br>● Most of the time<br>● All of the time | Yes, if most of the time or All of the time, no, otherwise | 1 if yes, otherwise,0 |
| understand_problems | How often do you have someone who understands your problems? | ● None of the time<br>● A little of the time<br>● Some of the time<br>● Most of the time<br>● All of the time | Yes, if most of the time or all of the time, no, otherwise | 1 if yes, otherwise,0 |
| poor_service | How often do you receive poorer service than others when you go to a doctor's office or other health care provider? | ● Almost every day<br>● At least once a week<br>● A few times a month<br>● A few times a year<br>● Less than once a year<br>● Never | Yes, if almost every day or at least once a week, no, otherwise | 0 if yes, otherwise 1 |
| doctor_afraid_of you | How often does a doctor or nurse act as if he or she is afraid of you when you go to a doctor's office or other health care provider? | ● Never<br>● Rarely<br>● Sometimes<br>● Most of the time<br>● Always | Yes, if most of the time or always, no, otherwise | 0 if yes, otherwise 1 |
| doctor_better_thanyou | How often does a doctor or nurse act as if he or she is better than you when you go to | ● Never<br>● Rarely<br>● Sometimes | Yes, if most of the time or always, no, otherwise | 0 if yes, otherwise 1 |

| | | | | |
|---|---|---|---|---|
| | a doctor's office or other health care provider?", | ● Most of the time<br>● Always | | |
| doctor_not_smart | How often does a doctor or nurse act as if he or she thinks you are not smart when you go to a doctor's office or other health care provider? | ● Never<br>● Rarely<br>● Sometimes<br>● Most of the time<br>● Always | Yes, if most of the time or always, no, otherwise | 0 if yes, otherwise 1 |
| move_home | In the last 12 months, how many times have you or your family moved from one home to another? Number of moves in past 12 months | 0 to n | Yes, if n, no if no move | 0 if yes, otherwise 1 |
| control_irritation | In the last month, how often have you been able to control irritations in your life? | ● Never<br>● Almost Never<br>● Sometimes<br>● Fairly Often<br>● Very Often | Yes, if Fairly Often or Very Often, no, otherwise | 1 if yes, otherwise 0 |
| angered_control | In the last month, how often have you been angered because of things that were outside of your control? | ● Never<br>● Almost Never<br>● Sometimes<br>● Fairly Often<br>● Very Often | Yes, if Fairly Often or Very Often, no, otherwise | 0 if yes, otherwise 1 |
| upset_unexpectdly | In the last month, how often have you been upset because of something that happened unexpectedly? | ● Never<br>● Almost Never<br>● Sometimes<br>● Fairly Often<br>● Very Often | Yes, if fairly often or very often, no, otherwise | 0 if yes, otherwise 1 |
| ability_handle | In the last month, how often have you felt confident about your ability to handle your personal problems? | ● Never<br>● Almost Never<br>● Sometimes<br>● Fairly Often<br>● Very Often | Yes, if Fairly Often or Very Often, no, otherwise | 1 if yes, otherwise 0 |
| piling_overcome | In the last month, how often have you felt difficulties were piling up so high that you could not overcome them? | ● Never<br>● Almost Never<br>● Sometimes | Yes, if Fairly Often or Very Often, no, otherwise | 0 if yes, otherwise 1 |

| | | | | |
|---|---|---|---|---|
| | | ● Fairly Often<br>● Very Often | | |
| stressed | In the last month, how often have you felt nervous and "stressed"?', | ● Never<br>● Almost Never<br>● Sometimes<br>● Fairly Often<br>● Very Often | Yes, if Fairly Often or Very Often, no, otherwise | 0 if yes, otherwise 1 |
| things_goingwell | In the last month, how often have you felt that things were going your way? | ● Never<br>● Almost Never<br>● Sometimes<br>● Fairly Often<br>● Very Often | Yes, if Fairly Often or Very Often, no, otherwise | 1 if yes, otherwise 0 |
| top_things | In the last month, how often have you felt that you were on top of things? | ● Never<br>● Almost Never<br>● Sometimes<br>● Fairly Often<br>● Very Often | Yes, if Fairly Often or Very Often, no, otherwise | 1 if yes, otherwise 0 |
| unable_controlthing | In the last month, how often have you felt that you were unable to control the important things in your life? | ● Never<br>● Almost Never<br>● Sometimes<br>● Fairly Often<br>● Very Often | Yes, if Fairly Often or Very Often, no, otherwise | 0 if yes, otherwise 1 |
| cope_things | In the last month, how often have you found that you could not cope with all the things that you had to do? | ● Never<br>● Almost Never<br>● Sometimes<br>● Fairly Often<br>● Very Often | Yes, if Fairly Often or Very Often, no, otherwise | 0 if yes, otherwise 1 |
| insulted | In your day-to-day life, how often are you called names or insulted? | ● Almost every day<br>● At least once a week<br>● A few times a month<br>● A few times a year<br>● Less than once a year<br>● Never | Yes, almost every day or at least once a week, no, otherwise | 0 if yes, otherwise 1 |

| | | | | |
|---|---|---|---|---|
| harassed | In your day-to-day life, how often are you threatened or harassed? | ● Almost every day<br>● At least once a week<br>● A few times a month<br>● A few times a year<br>● Less than once a year<br>● Never | Yes, almost every day<br>or at least once a week, no, otherwise | 0 if yes, otherwise 1 |
| less_courtesy_people | In your day-to-day life, how often are you treated with less courtesy than other people? | ● Never<br>● Rarely<br>● Sometimes<br>● Most of the time<br>● Always | Yes, if most of the time or always, no, otherwise | 0 if yes, otherwise 1 |
| less_respect_people | In your day-to-day life, how often are you treated with less respect than other people? | ● Never<br>● Rarely<br>● Sometimes<br>● Most of the time<br>● Always | Yes, if most of the time or always, no, otherwise | 0 if yes, otherwise 1 |
| people_afraidof you | In your day-to-day life, how often do people act as if they are afraid of you? | ● Never<br>● Rarely<br>● Sometimes<br>● Most of the time<br>● Always | Yes, if most of the time or always, no, otherwise | 0 if yes, otherwise 1 |
| dishonest_you | In your day-to-day life, how often do people act as if they think you are dishonest? | ● Almost every day<br>● At least once a week<br>● A few times a month<br>● A few times a year<br>● Less than once a year<br>● Never | yes, if almost every day or<br>At least once a week, no, otherwise | 0 if yes, otherwise 1 |
| people_smart | In your day-to-day life, how often do people act as if they think you are not smart? | ● Almost every day<br>● At least once a week<br>● A few times a month<br>● A few times a year<br>● Less than once a year<br>● Never | yes, if almost every day or<br>At least once a week, no, otherwise | 0 if yes, otherwise 1 |

| | | | | |
|---|---|---|---|---|
| people_better_thanyou | In your day-to-day life, how often do people act as if they're better than you are? | ● Almost every day<br>● At least once a week<br>● A few times a month<br>● A few times a year<br>● Less than once a year<br>● Never | yes, if almost every day or At least once a week, no, otherwise | 0 if yes, otherwise 1 |
| poor_restaurant | In your day-to-day life, how often do you receive poorer service than other people at restaurants or stores? | ● Almost every day<br>● At least once a week<br>● A few times a month<br>● A few times a year<br>● Less than once a year<br>● Never | yes, if almost every day or At least once a week, no, otherwise | 0 if yes, otherwise 1 |
| walk_transit | It is within a 10-15 minute walk to a transit stop (such as bus, train, trolley, or tram) from my home. Would you say that you... | ● Strongly disagree<br>● Somewhat disagree<br>● Somewhat agree<br>● Strongly agree<br>● Don't know/Not sure | yes, if somewhat agree or strongly agree, no, otherwise | 1 if yes, otherwise 0 |
| walking_distance | Many shops, stores, markets or other places to buy things I need are within easy walking distance of my home. Would you say that you... | ● Strongly disagree<br>● Somewhat disagree<br>● Somewhat agree<br>● Strongly agree<br>● Don't know/Not sure | yes, if somewhat agree or strongly agree, no, otherwise | 1 if yes, otherwise 0 |
| recreation_facility | My neighborhood has several free or low-cost recreation facilities, such as parks, walking trails, bike paths, recreation centers, playgrounds, public swimming pools, etc. Would you say that you... | ● Strongly disagree<br>● Somewhat disagree<br>● Somewhat agree<br>● Strongly agree<br>● Don't know/Not sure | yes, if somewhat agree or strongly agree, no, otherwise | 1 if yes, otherwise 0 |
| night_walk | The crime rate in my neighborhood makes it unsafe to go on walks at night. Would you say that you...', | ● Strongly disagree<br>● Somewhat disagree<br>● Somewhat agree<br>● Strongly agree<br>● Don't know/Not sure | yes, if somewhat agree or strongly agree, no, otherwise | 0 if yes, otherwise 1 |

| | | | | |
|---|---|---|---|---|
| day_walk | The crime rate in my neighborhood makes it unsafe to go on walks during the day. Would you say that you... | ● Strongly disagree<br>● Somewhat disagree<br>● Somewhat agree<br>● Strongly agree<br>● Don't know/Not sure | yes, if somewhat agree or strongly agree, no, otherwise | 0 if yes, otherwise 1 |
| speak_english | Since you speak a language other than English at home, we are interested in your own thoughts about how well you think you speak English. Would you say you speak English... | ○ Very well<br>○ Well<br>Not well<br>○ Not at all<br>○ Prefer not to answer<br>○ Don't know<br>● No<br>● Prefer not to answer | Yes, if very Well or well, no, otherwise | 1 if yes, otherwise 0 |
| bicycles | There are facilities to bicycle in or near my neighborhood, such as special lanes, separate paths or trails, or shared use paths for cycles and pedestrians. Would you say that you...', | ● Strongly disagree<br>● Somewhat disagree<br>● Somewhat agree<br>● Strongly agree<br>● Don't know/Not sure | yes, if somewhat agree or strongly agree, no, otherwise | 1 if yes, otherwise 0 |
| sidewalks | sidewalks= 'There are sidewalks on most of the streets in my neighborhood. Would you say that you...', | ● Strongly disagree<br>● Somewhat disagree<br>● Somewhat agree<br>● Strongly agree<br>● Don't know/Not sure | yes, if somewhat agree or strongly agree, no, otherwise | 0 if yes, otherwise 1 |
| place_live | Think about the place you live. Do you have problems with any of the following? Select all that apply | Bug infestation<br>● Mold<br>● Lead paint or pipes<br>● Inadequate heat<br>● Oven or stove not working<br>● No or not working smoke detector<br>● Water leaks<br>● None of the above | Yes, if present, no, otherwise | 0 if yes, otherwise 1 |

| | | | | |
|---|---|---|---|---|
| housing | What is the main type of housing in your neighborhood? | ● single family house, townhouses, or apartment | Yes, if single family house, townhouses, no, otherwise | 1 if yes, otherwise 0 |
| food_notlast | food_out='Within the past 12 months, were you worried whether your food would run out before you got money to buy more?') | ● Often true<br>● Sometimes true<br>● Never true | yes, if often true, no, otherwise | 0 if yes, otherwise 1 |
| | food_notlast= "Within the past 12 months, were you worried whether the food you had bought just didn't last and you didn't have money to get more?", | ● Often true<br>● Sometimes true<br>● Never true | yes, if often true, no, otherwise | 0 if yes, otherwise 1 |
| cannot_afford_prescription | Can't Afford Care: Prescription Medicines | No<br>Yes<br>PMI: Do not Know<br>PMI: Skip | Yes, if the response is "yes", otherwise no | 1 if yes, otherwise 0 |
| alternative_therapy | Alternative Therapies to Save Money | No<br>Yes<br>PMI: Do not Know<br>PMI: Skip | Yes, if the response is "yes", otherwise no | 1 if yes, otherwise 0 |
| skipp_medication | Skipped Med to Save Money | No<br>Yes<br>PMI: Do not Know<br>PMI: Skip | Yes, if the response is "yes", otherwise no | 1 if yes, otherwise 0 |
| low_cost | Lower Cost Rx to Save Money | No<br>Yes<br>PMI: Do not Know<br>PMI: Skip | Yes, if the response is "yes", otherwise no | 1 if yes, otherwise 0 |
| took_less_med | Took Less Med to Save Money | No<br>Yes<br>PMI: Do not Know<br>PMI: Skip | Yes, if the response is "yes", otherwise no | 1 if yes, otherwise 0 |
| another_country | Bought Rx from Other Country | No<br>Yes<br>PMI: Do not Know<br>PMI: Skip | Yes, if the response is "yes", otherwise no | 1 if yes, otherwise 0 |

| | | | | |
|---|---|---|---|---|
| delayed_filling | Delayed Filling Rx to Save Money | No<br>Yes<br>PMI: Do not Know<br>PMI: Skip | Yes, if the response ifs“yes”, otherwise no | 1 if yes, otherwise 0 |

*Include or classify all “PMI: Do not Know” or “PMI: Skip” responses from each question as “no”.

Figure S1: The domains of SDH reported in All of Us dataset

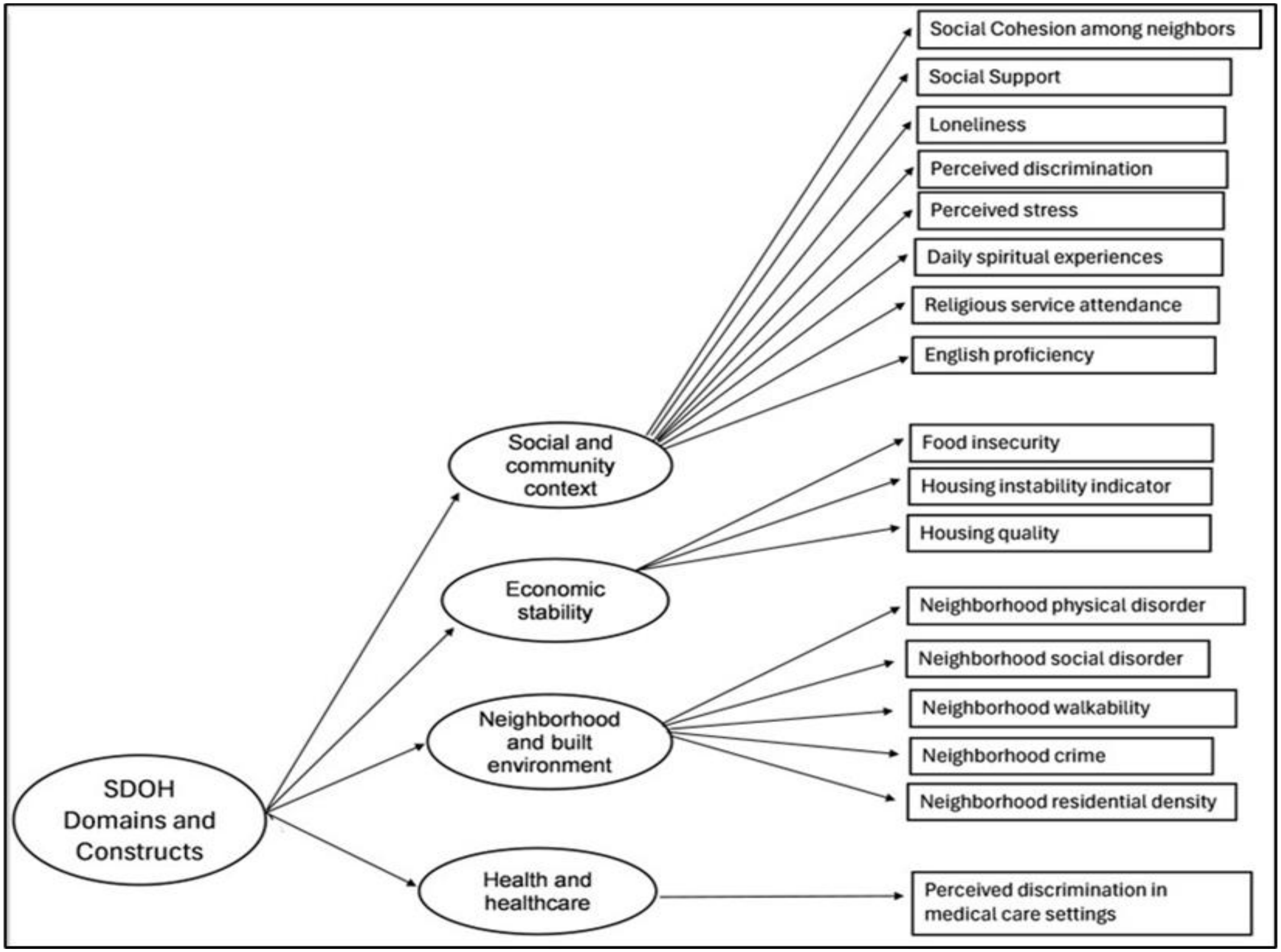

Table S3: Association between SDH features and CRMNA

| Characteristics | Overall CRMNA | Skip Medication | Alternative Therapy | Low Dose Medication | Cannot Afford Medication | Low cost Medication | Bought from Another Country | Delayed Filling |
|---|---|---|---|---|---|---|---|---|
| | AOR | AOR | AOR | AOR | AOR | AOR | AOR | AOR |
| **Gender** | | | | | | | | |
| Female | 1.29 [1.25- 1.33] | 1.40[1.31-1.49] | 1.58[1.49-1.68] | 1.45[1.36-1.54] | 1.37[1.31-1.45] | 1.29[1.25-1.34] | 0.83[0.77 0.89] | 1.54 1.46 1.62 |
| Other | 1.39 [1.29- 1.50] | 1.67[1.46-1.90] | 1.64[1.45-1.85] | 1.80[1.59-2.03] | 1.34[1.20-1.49] | 1.41[1.29-1.54] | 0.73[0.60-0.89] | 1.83 1.64 2.04 |
| Race | | | | | | | | |
| Black | 0.86 [0.81- 0.90] | 0.91[0.83-1.00] | 0.84[0.76-0.92] | 0.84[0.77-0.92] | 1.48[1.38-1.59] | 0.69[0.64-0.73] | 0.40[0.33-0.49] | 0.93 0.86 1.00 |
| Asian | 0.85 [0.77- 0.92] | 0.65[0.53-0.80] | 0.98[0.84-1.13] | 0.76[0.63-0.90] | 0.67[0.57-0.79] | 0.80[0.72-0.89] | 1.06[0.87-1.28] | 0.61 0.52 0.71 |
| Others | 0.90 [0.84-0.97] | 0.88[0.77-1.00] | 0.99[0.88-1.11] | 0.87[0.77-0.98] | 1.10[0.99-1.22] | 0.81[0.74-0.88] | 1.08[0.91-1.29] | 0.83[0.75- 0.93] |
| **Ethnicity** | | | | | | | | |
| Hispanic | 1.00 [0.93-1.08] | 0.93[0.82-1.06] | 1.06[0.94-1.19] | 0.87[0.77-0.99] | 1.09[0.98-1.21] | 0.90[0.83-0.98] | 1.29[1.09-1.54] | 0.91 0.81 1.01 |
| **Age** | | | | | | | | |
| ≥40-64 | 1.10 [1.06-1.14] | [1.20-1.12-1.28] | 0.89[0.84-0.94] | 1.17[1.10-1.25] | 1.08[1.02-1.14] | 1.30[1.25-1.36] | 1.03[0.93-1.14] | 1.11 1.05 1.17 |
| ≥65-74 | 1.07 [1.03- 1.12] | 0.97[0.89-1.06] | 0.51[0.47-0.56] | 0.95[0.88-1.03] | 0.91[0.85-0.97] | 1.43[1.36-1.51] | 1.33[1.19-1.48] | 0.84 0.79 0.90 |
| ≥75-84 | 0.92 [0.87- 0.97] | 0.66[0.57-0.75] | 0.29[0.25-0.33] | 0.70[0.62-0.79] | 0.59[0.53-0.65] | 1.21[1.14-1.29] | 1.55[1.36-1.76] | 0.52 0.47 0.58 |
| ≥85 | 0.67 [0.58- 0.77] | 0.36[0.22-0.55] | 0.30[0.20-0.43] | 0.42[0.28-0.61] | 0.39[0.28-0.54] | 0.86[0.72-1.01] | 1.09[0.79-1.47] | 0.30 0.21 0.43 |
| SDH3 | 1.16 1.12 1.20 | 1.38[1.28-1.48] | 1.17[1.10-1.24] | 1.34[1.25-1.43] | 1.33[1.26-1.41] | 0.94[0.90-0.98] | - | 1.33 1.26 1.41 |
| SDH7 | 1.36[1.32 -1.40] | - | 1.47[1.39-1.55] | - | - | 0.74[0.71-0.76] | - | 1.40 1.33 1.46 |
| SDH4 | 1.20 [1.16- 1.24] | 1.26[1.18-1.34] | 1.22[1.15-1.29] | 1.27[1.20-1.35] | 1.23 [1.17-1.30] | 0.87[0.84-0.91] | - | 1.29 1.23 1.36 |
| SD9 | 1.05 [1.02- 1.09] | 1.16[1.08-1.23] | - | 1.13[1.06-1.20] | 1.08[1.02-1.13] | 0.95[0.92-0.99] | - | 1.02 0.97 1.08 |
| SDH6 | 1.16 [1.10-1.22] | 1.12[1.04-1.21] | 1.10[1.03-1.19] | 1.18[1.10-1.27] | 1.15[1.08-1.23] | 0.91[0.86-0.96] | - | 1.16 1.09 1.23 |
| SDH5 | 1.41 [1.36-1.46] | 1.55[1.46-1.65] | 1.59[1.51-1.68] | 1.50[1.42-1.59] | 1.41[1.35-1.48] | 0.79[0.76-0.83] | - | 1.47 1.40 1.54 |
| SDH2 | 1.23 [1.17- 1.29] | 1.37[1.27-1.48] | 1.22[1.14-1.32] | 1.35[1.26-1.45] | 1.26[1.18-1.34] | 0.85[0.80-0.89] | - | 1.29 1.21 1.37 |
| SDH1 | 2.29 [2.19 - 2.40] | 3.55[3.33-3.79] | 1.82[1.70-1.94] | 3.25[3.05-3.46] | 3.56[3.37-3.76] | 0.60[0.57-0.63] | 1.33[1.19-1.47] | 3.18 3.01 3.36 |
| SDH10 | 1.21 [1.09-1.33] | 1.43[1.28-1.59] | 1.12[0.99-1.26] | 1.34[1.21-1.49] | 1.46[1.32-1.61] | 0.86[0.78-0.95] | - | 1.32 1.19 1.46 |
| SDH8 | 1.59 [1.50-1.69] | 1.72[1.59-1.87] | 1.75[1.62-1.89] | 1.71[1.58-1.85] | 1.64[1.52-1.76] | 0.71[0.67-0.76] | - | 1.66 1.54 1.78 |
| SDH11 | - | 1.36[1.23-1.52] | 1.38[1.24-1.53] | 1.36[1.22-1.50] | 1.35[1.23-1.48] | - | - | - |
| SDH2 | - | - | - | - | - | - | 0.41[0.36-0.47] | - |
| SDH17 | - | - | - | - | - | - | 1.07[0.97-1.17] | - |
| SDH16 | - | - | - | - | - | - | 1.00[0.89-1.13] | - |
| SDH15 | - | - | - | - | - | - | 1.01[0.90-1.14] | - |
| SDH14 | - | - | - | - | - | - | 1.19[1.07-1.33] | - |
| SDH19 | - | - | - | - | - | - | 0.99[0.88-1.11] | - |
| SDH20 | - | - | - | - | - | - | 1.02[0.91-1.14] | - |
| SDH18 | - | - | - | - | - | - | 1.50[1.30-1.71] | - |
| SDH13 | - | - | - | - | - | - | 1.24[1.08-1.43] | - |

AOR: Adjusted odds ratio. The dashed line indicates that the SDH was not ranked among the top ten features and was therefore not included in the regression analysis for that specific CRMNA. Statistical significance was defined as a p-value less than 0.05. Male, white, non-Hispanic, and <40 were references.

Figure S2: ROC curves for predicting cost-related medication non-adherence

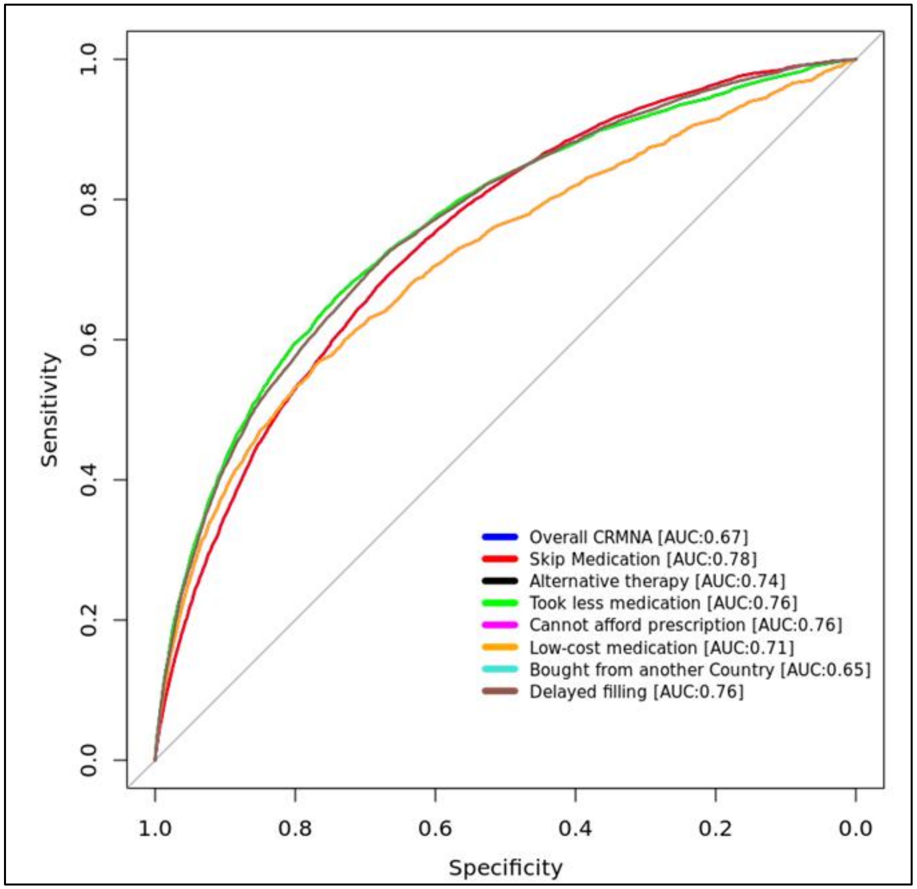
